# HIV care preferences among young people living with HIV in Lesotho: A secondary data analysis of the PEBRA cluster randomized trial

**DOI:** 10.1101/2022.07.29.22278205

**Authors:** Olivia Seiler, Mathebe Kopo, Mpho Kao, Thabo Ishmael Lejone, Nadine Tschumi, Tracy Renée Glass, Jennifer Anne Brown, Niklaus Daniel Labhardt, Alain Amstutz

## Abstract

Sub-Saharan Africa is home to 89% of all young people living with HIV, a key population with specific challenges and needs. In-depth knowledge of service demands is needed to tailor and differentiate service delivery for this group. We evaluated HIV care preferences among young people living with HIV who were part of the PEBRA (Peer Educator Based Refill of ART) cluster-randomized trial.

The PEBRA trial evaluated a novel model of care at 20 health facilities in Lesotho, Southern Africa. In the PEBRA model, a peer educator regularly assessed participant preferences regarding antiretroviral therapy (ART) refill location, SMS notifications (for adherence, drug refill, viral load) and general care support options, and delivered services accordingly over a 12-month period. We present these preferences, their changes over time and how often it was not feasible to deliver them.

At enrolment, 41 of 123 (33.3%) chose ART refill outside the health facility, compared to 8 of 123 (6.5%) after 12 months. Among those selecting clinic-based ART refill, many collected ART during the peer educator led Saturday clinic club, 45 of 123 (36.5%) at the beginning and 55 of 123 (44.7%) at the end. SMS reminder for adherence and/or ART refill was chosen by 51 of 123 (41.5%) at enrolment and 54 of 123 (44.7%) at the last assessment. Support by the peer educator was popular at the beginning (110 of 123 (89.4%)) and lower but still high at the end (85 of 123 (69.1%)). 13 of 123 (10.6%) participants chose support by the nurse only at the first and 21 of 123 (17.1%) at the last assessment. The overarching trial was prospectively registered on ClinicalTrials.gov (NCT03969030).

Our longitudinal preference assessment among young people living with HIV showed a sustained interest in SMS notifications for adherence and refill visits as well as in additional support by a peer educator. ART refill outside the health facility was not as popular as expected; instead, medication pick-up at the facility, especially during Saturday clinic clubs, was favoured.

## Introduction

According to the UNAIDS 2021 report, 2 out of 7 new HIV infections in 2019 occurred in young people aged 15 to 24 years and sub-Saharan Africa is home to 89% of all young people living with HIV [1,2]. This age group makes up a substantial part of the HIV-positive population in sub-Saharan Africa with a share of 20% [3]. AIDS-related deaths are the leading cause of mortality in this population [4].

In Lesotho, the Demographic and Health Survey of 2014 showed that 10% of young people were living with HIV with 13% of women and 6% of men affected [5]. Young people living with HIV are a vulnerable subpopulation that faces distinctive challenges and therefore needs special attention on the way to the goal of ending the AIDS epidemic by 2030 [4,6].

Differentiated Service Delivery (DSD) is an approach that shifts from a one-size-fits-all model to a patient-centered approach, with the idea to better meet the individual needs of people living with HIV [7]. Several DSD models have been implemented for adults living with HIV who have stable viral suppression, and research has been conducted looking at the sustainability and cost-effectiveness of such models as well as participants’ preferences [8–11]. A situational analysis from 2018 conducted by Paediatric-Adolescent Treatment Africa reported a lack of published literature about DSD models specific to young people [12]. To tailor programs according to the needs and demands of young people living with HIV, knowledge of their preferences is required.

In this secondary analysis, we used data collected in the intervention arm of the PEBRA (Peer educator Based Refill of ART) cluster randomized trial in rural Lesotho [13]. We evaluated participants’ HIV care preferences, their feasibility, and intra-individual changes of preferences throughout the 12-month study period.

## Materials and Methods

### Study design and participants

This is a pre-planned secondary analysis of data collected in the intervention arm of the PEBRA trial – a cluster randomized controlled trial conducted at 20 nurse-led health facilities in three districts of Lesotho between November 2019 and April 2021. The PEBRA trial assessed the effectiveness of a peer educator-coordinated preference-based antiretroviral therapy (ART) service delivery model among young people living with HIV in Lesotho (PEBRA model). PEBRA enrolled young people living with HIV aged 15-24 years taking ART. The 20 health facilities (clusters) were spread over three mostly rural districts in Lesotho: Leribe, Butha-Buthe and Mokhotlong. The clinics were randomized in a 1:1 allocation to an intervention (PEBRA model) and a control arm. In this study, we only included data of the 123 of the 150 participants at the 10 PEBRA model clinics who remained in care over the entire 12-month period. Detailed information about the setting, design, eligibility, randomization, data collection and management as well as about the primary and secondary endpoint are published in the PEBRA study protocol [13] and the main results are under review in a peer-reviewed journal (cite PEBRA).

### The PEBRA model

The PEBRA model was designed in collaboration with peer educators, young people living with HIV, youth advocates, clinical staff and application developers during several workshops supported and coordinated by two local non-profit organizations (SolidarMed & Sentebale) as well as the Ministry of Health of Lesotho. It consists of three pillars: ART refill location, SMS notifications and general care support and made use of preexisting structures at the local clinics. For the ART refill location, participants could choose from the following options: refill at the clinic with the option of pick-up within the Saturday clinic club, refill at the village health worker’s home, home delivery by the peer educator, refill at the community adherence club or refill by a treatment buddy (Supplementary Table 1). Regarding SMS notifications, the participants could choose to get a notification reminding them to take their ART (adherence reminder), to remind them of the next ART refill (refill reminder) and to receive a viral load result message (viral load result notification) (Supplementary Table 2). It was possible to opt in to more than one notification option and, for each notification, they could specify the message content, time, and frequency. The third pillar of the PEBRA model is the additional support that participants could choose from. The different possibilities were: support through the nurse at the clinic, Saturday clinic club, community youth club, phone call by peer educator, home-visit by the peer educator, school visit and health talk by peer educator, pitso (a community gathering) visit and health talk by peer educator, condom demonstration, more information about contraceptives, more information about voluntary male medical circumcision (VMMC), linkage to young mothers group (for pregnant women), linkage to a female social asset building model, and more information about gender-based violence/legal aid. It was possible for participants to choose multiple sources of support. Each option is explained in more detail in Supplementary Table 3.

At each of the 10 intervention facilities, a trained peer educator delivered the PEBRA model using the PEBRApp, a tablet-based application designed specifically for the PEBRA study. The peer educator conducted a preference assessment among his/her participants every three months or every month for virally supressed or unsuppressed (> 999 copies/mL) participants, respectively. At every preference assessment visit, all the options were shown to the participants visually in the PEBRApp and explained individually. Subsequently, participants were asked which options they preferred. Then, the peer educator assessed if the chosen options were feasible (e.g. not every community had an established community youth club or the participants lived too far from the peer educator’s home, etc.) and if not feasible, the second-best option was chosen and delivered. The PEBRApp helped the peer educator to keep in regular contact with participants and keep track of participants’ preferences and ART refills. Together with the nurses and other clinic staff, the peer educator delivered services according to preferences and feasibility. The chosen SMS notifications were sent out automatically from the PEBRApp including a call-back option to the peer educators number.

### Variables and time points of interest

We included preference data for all three pillars of the PEBRA model over the course of the 12-month trial period. The main variables of interest for this analysis were the proportion of participants requesting alternative ART refill than individual pick-up at the clinic, adherence and refill reminder notifications, and additional support options provided by the peer educator. We assessed the feasibility of providing selected options during all PEBRA preference assessments. For the longitudinal assessments in preferences over the study period, three time points were considered: 1) enrolment, 2) 6 months after enrolment (range: 5-7 months), and 3) 12 months after enrolment (range: 11-14 months). We chose these three timepoints following the SPIRIT diagram of the PEBRA trial [13].

### Sankey diagrams

We created Sankey diagrams using the three defined timepoints and grouping preferences within each pillar of the PEBRA model. ART refill locations are shown in the categories of inquiry. SMS notifications were grouped as: adherence and / or refill reminders, which are not available in standard care; only viral load notifications or no notifications; and no cell phone available (see also Supplementary Table 2). Support options were grouped as peer educator support; nurse support; and other support (see also Supplementary Table 3). Peer educator support included Saturday clinic club (monthly gathering, led by the peer educator), community youth club, a phone call by the peer educator, a home visit by the peer educator, a school visit and health talk by the peer educator, or a pitso visit and health talk by the peer educator. These options were developed specifically for the PEBRA model and are not otherwise available. Nurse support corresponded to the usual standard of care. “Other” support options included support that was a one-time support on the day of the assessment such as condom demonstrations, information about contraceptives, information about VMMC, linkage to young mothers’ groups (for pregnant participants; DREAMS or Mothers-to-Mothers), linkage to a female social asset building model (for female participants; WORTH) and information about legal aid and gender-based violence. These “other” support options could be provided either by the peer educator, the nurse, or other staff at the health facility.

### Statistical analyses and software

We used absolute and relative frequencies to describe categorical data and medians and interquartile ranges for continuous variables.

The data analysis was performed in R (Version: R 4.1.1 GUI 1.77 High Sierra build). The Sankey Diagrams were built with SankeyMATIC (https://sankeymatic.com/build/).

### Ethical statement

The protocol of the PEBRA trial was approved by the National Health Research and Ethics Committee of the Ministry of Health of Lesotho (118–2019; June 03, 2019) and the ethics committee in Switzerland (Ethikkommission Nordwest-und Zentralschweiz; 2019-00480; June 14, 2019). The trained study staff obtained the individual written informed consent from the participants before inclusion into the PEBRA trial. As outlined in the approved PEBRA study protocol [13], in order to minimize selection bias, the ethics committees agreed to waive parental consent for the 15-17 years old study participants. Illiterate study participants provided a thumbprint and a literate witness (independent to the trial and chosen by the participant) co-signed the form. The informed consent was provided in the local language, Sesotho, and the participant received a copy of the consent form. The PEBRA trial was prospectively registered on clinicaltrials.gov (NCT03969030. Registered on 31 May 2019).

## Results

### Characteristics of the study population

The PEBRA model group enrolled 150 participants, of whom 123 were in care at 12 months and only these were included in the analysis of this manuscript. Detailed socio-demographic and clinical characteristics including viral loads disaggregated by gender can be found in the supplements of this paper (Supplementary Tables 4-6) and are part of the overall baseline characteristics table in the PEBRA main publication (under review) (cite PEBRA) In brief, the median age was 18.2 [interquartile range (IQR) 16.5 - 21.8] years, 79 of 123 (64.2%) were female, 121 of 123 (98.4%) were heterosexual and the median number of completed school years was 9.0 [IQR 7.0 - 10.0]. Asked about their occupation, 52 (42.3%) answered that they were attending school, 9 (7.3%) that they were (self-) employed, and the remaining 62 of 123 (50.4%) stated that they did not have an occupation. Of the 123 participants, 91 (74.0%) were single, 29 (23.6%) were married, 2 (1.6%) were separated and 1 (0.8%) was widowed. At the time of enrolment, 32 of 123 (26.0%) participants had one or more children and among women, 14 of 79 (17.7%) were pregnant or breastfeeding. The median number of years since HIV diagnosis was 5.8 [IQR 3.1 - 11.1], and the median number of years since starting ART was 5.0 [IQR 3.0 - 9.3]. None of the participants was on tuberculosis treatment. At baseline, 68 of 123 (55.3%) had a documented viral load < 20 copies/mL, 27 of 123 (22.0%) had a viral load of 20-999 copies/mL, and 14 of 123 (11.4%) participants had a viral load >999 copies/mL; the remaining 14 of 123 (11.4%) didn’t have a documented viral load result.

### ART refill preferences and changes over time

We assessed changes in preferences over the 12-month study period for the three pillars of PEBRA: ART refill options, messages, and support options (Figure 1-3). We report here only preferences that were eventually also carried out. The number of service preference that were not feasible to deliver are report in chapter XXX below.

**Figure 1.**
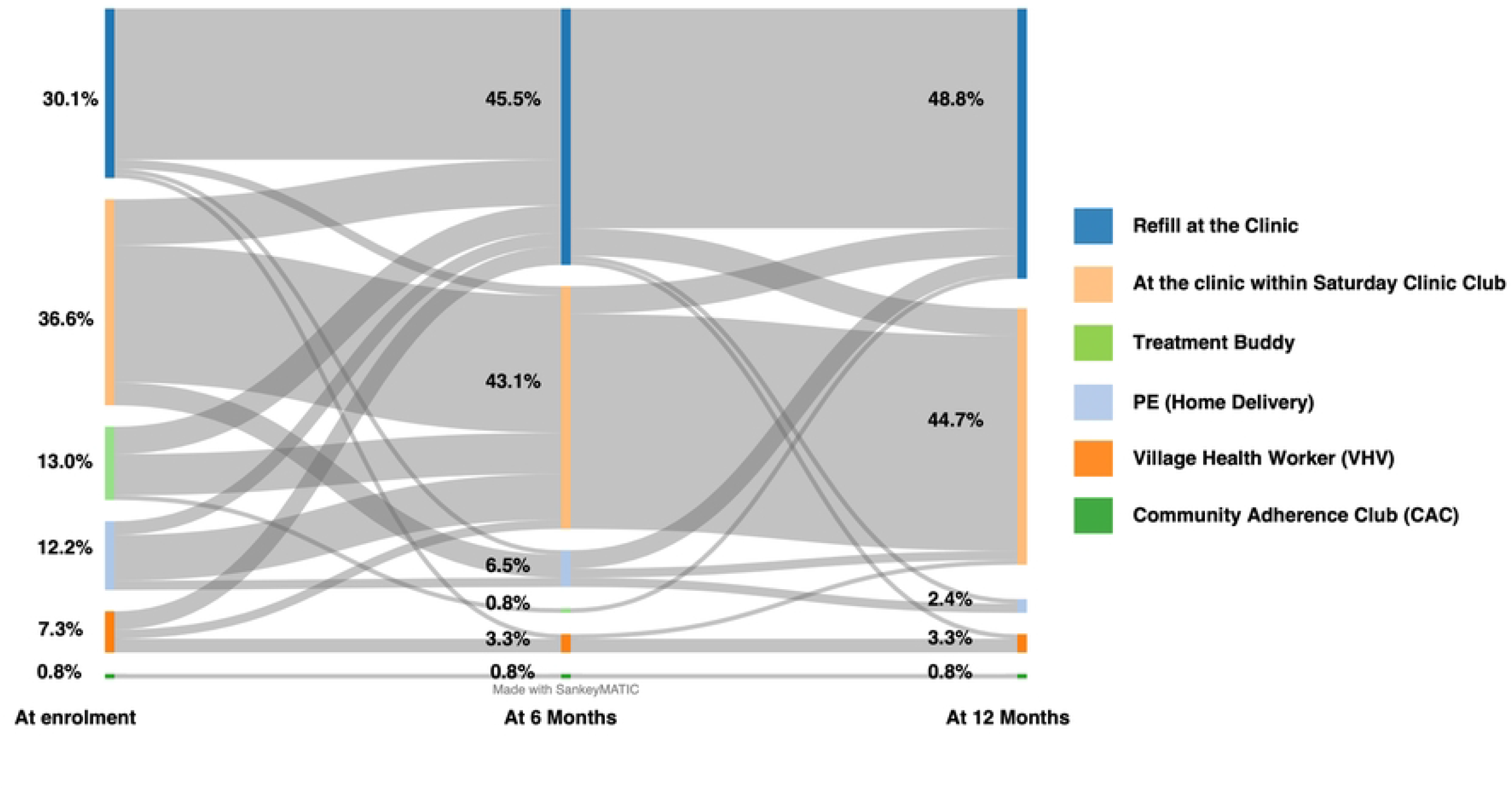
Sankey diagram for longitudinal visualization of ART refill site preferences

At enrolment, 41 of 123 (33.3%) intervention participants made use of the offer for an alternative ART refill option (Figure 1, Table 1). ART pick up by a treatment buddy was chosen by 16 of 123 (13.0%) participants, and 15 of 123 (12.2%) wanted to get their medication delivered to their home by the peer educator. The village health workers home was the preferred refill site for 9 of 123 (7.3%) participants and 1 of 123 participants wanted to pick up the ART in the community adherence club. Out of the 82 of 123 (66.7%) participants who chose to pick up their medication in the clinic, 45 of 123 (36.1%) did so within the Saturday clinic club.

**Table 1.** ART refill site preferences by gender

|  | First assessment | Middle assessment | Last assessment |
| --- | --- | --- | --- |

| level | Overall | female | male | Overall | female | male | Overall | female | male |
| --- | --- | --- | --- | --- | --- | --- | --- | --- | --- |
| n | 123 | 79 | 44 | 123 | 79 | 44 | 123 | 79 | 44 |
| At the clinic (%) | 37 (30.1) | 29 (36.7) | 8 (18.2) | 56 (45.5) | 43 (54.4) | 13 (29.5) | 60 (48.8) | 48 (60.8) | 12 (27.3) |
| At the clinic within the SCC (%) | 45 (36.6) | 23 (29.1) | 22 (50) | 53 (43.1) | 28 (35.4) | 25 (56.8) | 55 (44.7) | 27 (34.2) | 28 (63.6) |
| Treatment Buddy (%) | 16 (13.0) | 9 (11.4) | 7 (15.9) | 1 (0.8) | 1 (1.3) | 0 (0) | 0 (0.0) | 0 (0) | 0 (0) |
| peer educator (home delivery) (%) | 15 (12.2) | 9 (11.4) | 6 (13.6) | 8 (6.5) | 3 (3.8) | 5 (11.4) | 3 (2.4) | 1 (1.3) | 2 (4.5) |
| VHW (at the VHW's home) (%) | 9 (7.3) | 8 (10.1) | 1 (2.3) | 4 (3.3) | 3 (3.8) | 1 (2.3) | 4 (3.2) | 2 (2.5) | 2 (4.5) |
| CAC (Community Adherence Club) (%) | 1 (0.8) | 1 (1.3) | 0 (0) | 1 (0.8) | 1 (1.3) | 0 (0) | 1 (0.8) | 1 (1.3) | 0 (0) |

At the end, the proportion of the outside clinic refill preferences had shrunk to 8 of 123 (6.5%). Of which 3 of 123 (2.4%) chose home delivery by the peer educator, another 4 of 123 (3.3%) the Village health workers home and 1 of 123 (0.8%) wanted to pick the ART refill up at the community adherence club. The remaining 114 (93.5%) preferred to pick up the medication at the health facility, with about half of these 55 of 123 (44.7%) refilling ART at a Saturday clinic club.

### SMS notification preferences and changes over time

Figure 2 and table 2 summarize the SMS notification preferences over time. At enrolment 72 of 123 (58.5%) had access to a cell phone where they could receive confidential information on, this number increased to 80 of 123 (65.0%) at the end. The number of participants who wished to receive either a refill or an adherence SMS reminder was 51 of 123 (41.5%) at enrolment and 54 of 123 (44.7%) at the last assessment. The option to receive only VL notifications or no notifications was chosen by 21 of 123 (17.0%) participants at the beginning of the study respectively by 25 of 123 (20.3%) at the end.

**Figure 2.**
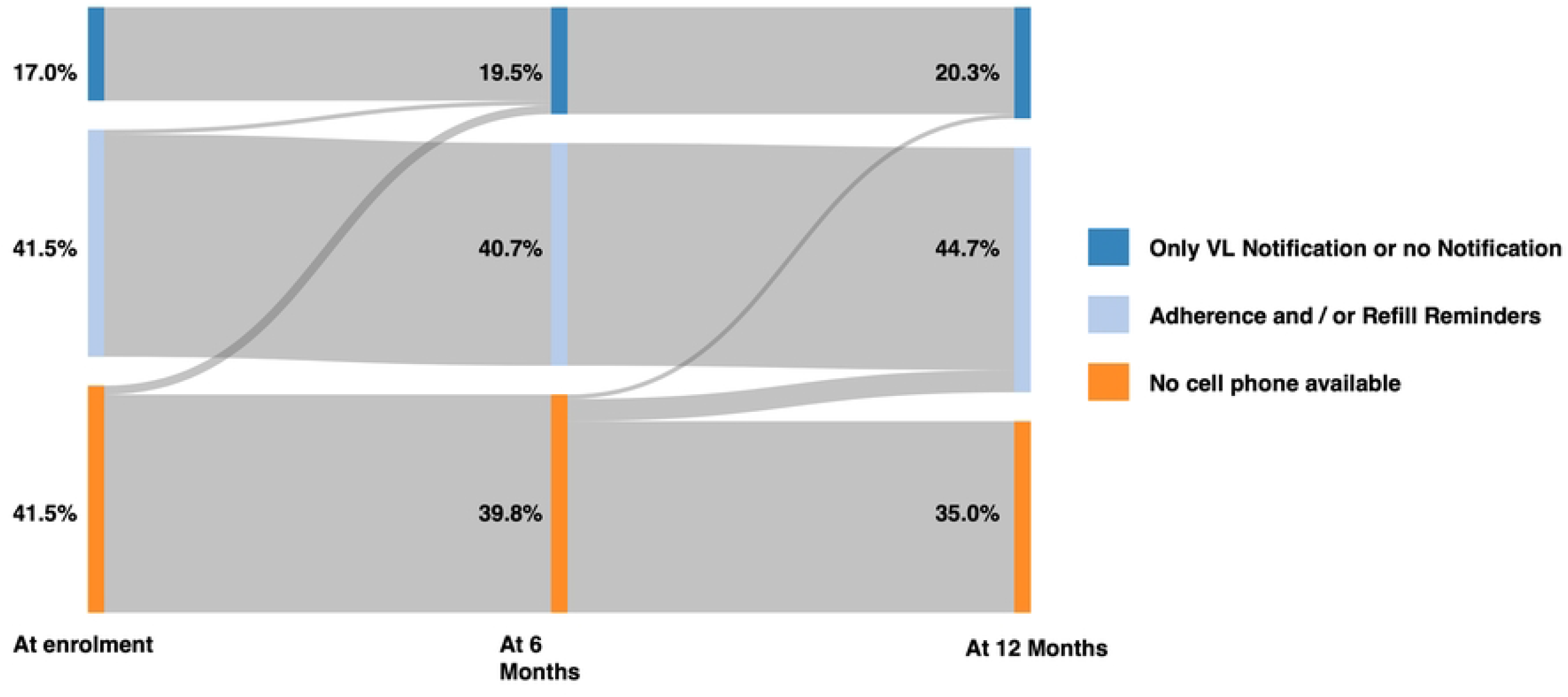
Sankey diagram for longitudinal visualization of SMS notification preferences

**Table 2.** SMS reminder preferences by gender

|  |  | First assessment |  |  | Middle assessment |  |  | Last assessment |  |  |
| --- | --- | --- | --- | --- | --- | --- | --- | --- | --- | --- |
|  | level | Overall | female | male | Overall | female | male | Overall | female | male |
| n |  | 123 | 79 | 44 | 123 | 79 | 44 | 123 | 79 | 44 |
| Cell phone available | No | 51 (41.5) | 27 (34.2) | 24 (54.5) | 49 (39.8) | 24 (30.8) | 24 (54.5) | 43 (35.0) | 23 (29.1) | 20 (45.5) |
| (%) | Yes | 72<br>(58.5) | 52<br>(65.8) | 20<br>(45.5) | 74<br>(60.2) | 54<br>(69.2) | 20<br>(45.5) | 80<br>(65.0) | 56<br>(70.9) | 24<br>(54.5) |
| Adherence reminders chosen (%) | No | 33<br>(26.8) | 22<br>(27.8) | 11<br>(25.0) | 31<br>(25.2) | 21<br>(26.9) | 10<br>(22.7) | 35<br>(28.5) | 23<br>(29.1) | 12<br>(27.3) |
|  | Yes | 39<br>(31.7) | 30<br>(38.0) | 9<br>(20.5) | 43<br>(35.0) | 33<br>(42.3) | 10<br>(22.7) | 45<br>(36.6) | 33<br>(41.8) | 12<br>(27.3) |
| Refill reminders chosen (%) | No | 31<br>(25.2) | 22<br>(27.8) | 9<br>(20.5) | 31<br>(25.2) | 22<br>(28.2) | 9<br>(20.5) | 30<br>(24.4) | 20<br>(25.3) | 10<br>(22.7) |
|  | Yes | 41<br>(33.3) | 30<br>(38.0) | 11<br>(25.0) | 43<br>(35.0) | 32<br>(41.0) | 11<br>(25.0) | 50<br>(40.7) | 36<br>(45.6) | 14<br>(31.8) |
| Viral load notifications chosen (%) | No | 4 (3.3) | 3 (3.8) | 1<br>(2.3) | 13<br>(10.6) | 9<br>(11.5) | 4<br>(9.1) | 14<br>(11.4) | 9<br>(11.4) | 5<br>(11.4) |
|  | Yes | 68<br>(55.3) | 49<br>(62.0) | 19<br>(43.2) | 61<br>(49.6) | 45<br>(57.7) | 16<br>(36.4) | 66<br>(53.7) | 47<br>(59.5) | 19<br>(43.2) |

### Support options and changes over time

Support by the peer educator was chosen by 110 of 123 (89.4%) participants at the first timepoint and decreased to 85 of 123 (69.1%) by end of the study (Figure 3, Table 3). At enrolment there were 13 of 123 (10.6%) participants who chose only support by the nurse at the clinic. At the last time point, 21 of 123 (17.1%) participants chose only support by the nurse, 1 of 123 (0.8%) chose only support without nurse or peer educator involvement and 16 of 123 (13.0%) participants wanted to have no support at all.

**Figure 3.**
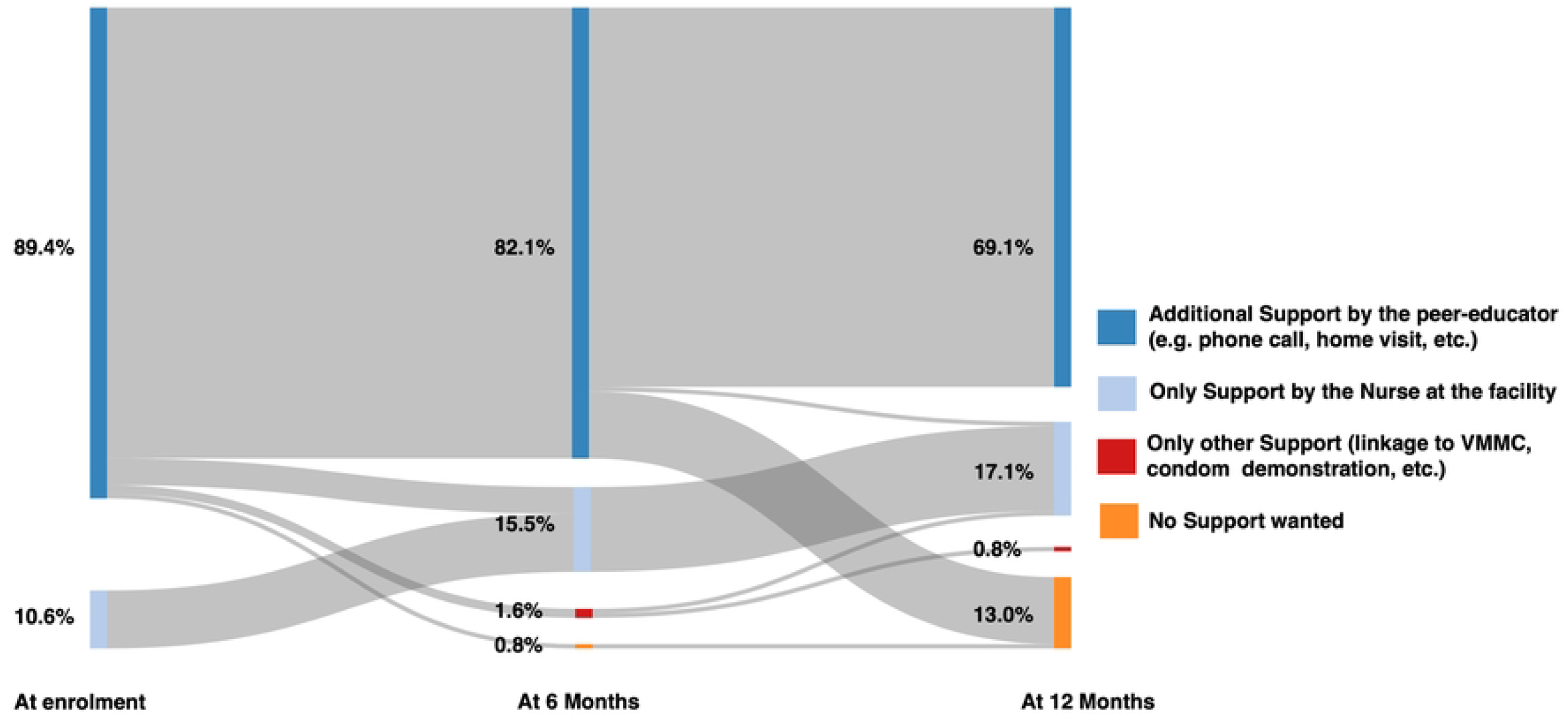
Sankey diagram for longitudinal visualization of support preferences

**Table 3.** Support preferences by gender

|  | First assessment |  |  | Middle assessment |  |  | Last assessment |  |  |
| --- | --- | --- | --- | --- | --- | --- | --- | --- | --- |
| level | Overall | female | male | Overall | female | male | Overall | female | male |
| n | 303 | 183 | 119 | 275 | 164 | 111 | 245 | 151 | 94 |
| *Peer educator support* |  |  |  |  |  |  |  |  |  |
| Saturday Clinic Club (SCC) (%) | 70<br>(23.1) | 39<br>(21.3) | 31<br>(26.1) | 56<br>(20.4) | 29<br>(17.7) | 27<br>(24.3) | 57<br>(23.3) | 28<br>(18.5) | 29<br>(30.9) |
| Community Youth Club (CYC) (%) | 2 (0.7) | 2 (1.1) | 0 (0.0) | 0 (0.0) | 0 (0.0) | 0 (0.0) | 0 (0.0) | 0 (0.0) | 0 (0.0) |
| Phone call by peer educator (%) | 60<br>(19.8) | 43<br>(23.5) | 16<br>(13.4) | 65<br>(23.6) | 43<br>(26.2) | 22<br>(19.8) | 68<br>(27.8) | 45<br>(29.8) | 23<br>(24.5) |
| Home-visit by peer educator (%) | 13 (4.3) | 6 (3.3) | 7 (5.9) | 14 (5.1) | 11<br>(6.7) | 3 (2.7) | 12 (4.9) | 7 (4.6) | 5 (5.3) |
| School visit and health talk by peer educator (%) | 5 (1.7) | 2 (1.1) | 3 (2.5) | 2 (0.7) | 2 (1.2) | 0 (0.0) | 0 (0.0) | 0 (0.0) | 0 (0.0) |
| Pitso visit and health talk by peer educator (%) | 2 (0.7) | 1 (0.5) | 1 (0.8) | 0 (0.0) | 0 (0.0) | 0 (0.0) | 0 (0.0) | 0 (0.0) | 0 (0.0) |
| *Nurse support* |  |  |  |  |  |  |  |  |  |
| By the nurse at the clinic (%) | 109 (36.0) | 70 (38.3) | 39 (32.8) | 98 (35.6) | 62 (37.8) | 36 (32.4) | 80 (32.7) | 52 (34.4) | 28 (29.8) |
| *Other support* |  |  |  |  |  |  |  |  |  |
| Condom demonstration (%) | 22 (7.3) | 11 (6.0) | 11 (9.2) | 15 (5.5) | 8 (4.9) | 7 (6.3) | 10 (4.1) | 8 (5.3) | 2 (2.1) |
| More information about contraceptives (%) | 10 (3.3) | 5 (2.7) | 5 (4.2) | 11 (4.0) | 5 (3.0) | 6 (5.4) | 1 (0.4) | 1 (0.7) | 0 (0.0) |
| More information about VMMC (%) | 5 (1.7) | NA | 5 (4.2) | 5 (1.8) | NA | 5 (4.5) | 0 (0.0) | NA | 0 (0.0) |
| For pregnant: Linkage to young mothers group (DREAMS or Mothers-to-Mothers) (%) | 1 (0.3) | 1 (0.5) | NA | 0 (0.0) | 0 (0.0) | NA | 0 (0.0) | 0 (0.0) | NA |
| For females: Linkage to a female WORTH group (Social Asset Building Model) (%) | 0 (0.0) | 0 (0.0) | NA | 0 (0.0) | 0 (0.0) | NA | 0 (0.0) | 0 (0.0) | NA |
| More information about legal aid and gender-based violence (%) | 4 (1.3) | 3 (1.6) | 1 (0.8) | 8 (2.9) | 4 (2.4) | 4 (3.6) | 1 (0.4) | 1 (0.7) | 0 (0.0) |
| *No support* |  |  |  |  |  |  |  |  |  |
| No support wanted (%) | 0 (0.0) | 0 (0.0) | 0 (0.0) | 1 (0.4) | 0 (0.0) | 1 (0.9) | 16 (6.5) | 9 (6.0) | 7 (7.4) |

### Overall preferences and feasibility

During the entire study period, the peer educators conducted a total of 800 preference assessments among the 123 participants. The median number of assessments per participant was 6 [IQR 5 - 6].

ART refill at the clinic was chosen in 671 of 800 (83.9%) and refill via a treatment buddy in 35 of 800 (4.4%) assessments. In all other instances, a refill outside the clinic was the first choice. However, among the 53 cases where the peer educator-delivery option was first choice, 12 (22.6%) had to be changed to another refill option due to feasibility constraints.

During the 800 preference assessments, a total of 1037 different SMS notifications were chosen: 435 of 1037 (41.9%) were VL notifications, 304 (29.3%) were refill reminders, and 298 (28.7%) were adherence reminders (Supplementary Table 8). Among the adherence reminders, most (257 of 298; 86.2%) were daily messages, followed by weekly (27 of 298; 9.1%) and monthly (14 of 298; 4.7%) messages (Supplementary Table 7). Most participants chose to receive the adherence reminders in the morning (6-10AM; 130 of 298; 43.8%) or evening (7PM-0AM; 134 of 298; 45.0%).

Among the 1839 support options chosen in the 800 assessments, peer educator support, nurse support, other support, and no support were selected 1014 (55.2%), 622 (33.8%), 168 (9.1%), and 35 (1.9%) times, respectively. As with the refill options, chosen support options were not always feasible. In 27 instances, a home visit by the peer educator was not feasible due to distance, and in six instances, a pitso visit was not possible for the same reason. Community youth clubs and Saturday clinic clubs were selected but not available in the participant’s community in 39 and at the participants’ clinic in eight instances, respectively. In 3 cases, linkage to a female WORTH group wasn’t feasible. In total, 83 support choices couldn’t be delivered; this was 4.5% of all care support demanded.

## Discussion

HIV care preference data among young people taking ART at HIV clinics in rural Lesotho revealed, that ART refill outside the clinic was not as popular as expected. At enrolment, 33.3% chose ART refill outside the health facility, however, only six months later, close to 90% were choosing to get refills at the clinic despite little issues of feasibility. Furthermore, the data revealed that SMS notifications such as adherence and refill reminders were widely chosen throughout the one-year study period. Additional support by the peer educator was feasible and highly popular when the study was introduced (89.4%), decreased over the 12-month period, but remained popular towards the end (69.1%).

Despite the high interest and urgency statements by the WHO and other international organisations and the knowledge gaps regarding this population, the pace of interventional adolescent HIV research is still slow [2,14]. There is some research with promising findings regarding DSD models for young people in Africa [15–17]. A small number of studies on HIV pre-exposure prophylaxis preferences among young people exist [18,19], but none assessing service or care preferences among young people living with HIV. In a formative study about engagement of young people in sexual and reproductive health, there was great interest in accessing health services at community hubs rather than the health facility [20], however, this finding does not coincide with our results.

SMS notifications and peer support were the alternatives with the highest uptake. Peer support generates trust and therefore hopefully stability in the long run [21,22]. However, it is important to note that 13% of participants wanted no support at all and 17% only support form the nurse at the health facility. Rather surprising was the high share of participants preferring to come to the clinic for their ART refill rather than the more decentralized options. The reason was not that the decentralized options were unfeasible (only 1.5% first choices were turned down due to feasibility). While decentralizing ART pick-up sounds appealing by removing structural barriers [9,23–25], we could not confirm this statement in practice among the study population. As the study was run during the COVID-19 pandemic where mobility and public life were reduced to a minimum, we would have expected more decentralized ART refills in this setting.

The Saturday clinic clubs may have played an important role for clinic refill as they are well established at most clinics. Also, nurses at the study facilities received a training in adolescent-friendly service delivery before the inception of the study and thus might have contributed to more clinic-based service choices. People living with HIV are still afraid of stigmatization [26–28]. This is one possible reason why study participants may have preferred to come to the clinic to pick up their medication, because it makes them feel less visible. It is also possible that the participants did not trust their peers or community members to provide ART refills or were dissatisfied with the services. The exact reasons remain to be explored in further research to adapt services to the needs of people living with HIV during the sensitive phase of adolescence and young adulthood.

This study has several limitations. The first concerns data collection. Despite training the peer educators on data collection and providing instructions on how to present the options to the participants, we cannot exclude the possibility of peer educators’ attitudes influencing participant preferences. Conditioning cannot be ruled out since the participants and peer educators knew that not all options were feasible for all participants at all times. Second, Lesotho has a unique geography, and the presented data originates from rural areas. Participants of urban areas of Lesotho may have different preferences. While it demonstrated the feasibility of and demand for alternative care options, these cannot be expected to lead to direct improvements in clinical outcomes.

## Conclusions

This longitudinal preference assessment among young people living with HIV in rural areas of Lesotho is a first of its kind. It shows that ART refill outside the health facility was not as popular as expected; instead, medication pick-up at the facility, especially during Saturday clinic clubs, was favoured. More research is needed to investigate the underlying reasons of each preference pattern. Overall, this key population has a clear interest in SMS notifications for adherence and upcoming refill visits as well as additional support provided by a peer educator.

## Data Availability

A key pseudo-anonymized individual participant dataset collected during the study, along with a data dictionary, will be made available at the time of publication through the data repository Zenodo with open access.

## Acknowledgements

We would like to recognize the hard work and valuable contributions of the peer educators in all three districts, the tireless support of the SolidarMed and Sentebale team as well as the District Health Management Teams. A special thanks goes to Tlotliso Mafantiri (Tech4All Lesotho) and Christoph Schwizer (www.christophschwizer.ch), who jointly developed the PEBRApp, as well as Ruben Dill for graphic designs. We thank the involved health facilities for their dedication to this project and we gratefully acknowledge the adolescents and young people living with HIV participating in this trial.

## Funding

This study was funded by the CIPHER grant from the International AIDS Society, obtained by AA.

## Notes

### Competing Interest Statement

The authors have declared no competing interest.

### Clinical Trial

It is registered with ClinicalTrials.gov (NCT03969030).

### Clinical Protocols

https://bmcpublichealth.biomedcentral.com/articles/10.1186/s12889-020-08535-6

### Funding Statement

The funders had no role in study design, data collection and analysis, decision to publish, or preparation of the manuscript.

### Author Declarations

The protocol of the PEBRA trial was approved by the National Health Research and Ethics Committee of the Ministry of Health of Lesotho (118–2019 June 03, 2019) and the ethics committee in Switzerland (Ethikkommission Nordwest- und Zentralschweiz 2019-00480 June 14, 2019).

